# Pregnancy and Neonatal Outcomes in SARS-CoV-2 Infection: a systematic review

**DOI:** 10.1101/2020.05.11.20098368

**Authors:** Reem S Chamseddine, Farah Wahbeh, Frank Chervenak, Laurent J Salomon, Baderledeen Ahmed, Arash Rafii

## Abstract

With the emergence of SARS-CoV-2 and its rapid spread, concerns regarding its effects on pregnancy outcomes have been growing. We reviewed 164 pregnancies complicated by maternal SARS-CoV-2 infection across 20 studies. The most common clinical presentations were fever (57.9%), cough (35.4%), fatigue (15.2%), and dyspnea (12.2%). Only 2.4% of patients developed respiratory distress. Of all patients, 84.5% delivered via Cesarean section, with a 23.9% rate of maternal gestational complications, 20.3% rate of preterm delivery, and a concerning 2.3% rate of stillbirth delivery. Relative to known viral infections, the prognosis for pregnant women with SARS-CoV-2 is good, even in the absence of specific antiviral treatment. However, neonates and acute patients, especially those with gestational or pre-existing co-morbidities, must be actively managed to prevent severe outcomes.

## Introduction

The emergence of SARS-CoV-2 as a novel infection in late December 2019 poses unique challenges to healthcare systems and practitioners. Chief among them is the management for pregnant women who are infected with SARS-CoV-2. Pregnant women are prone to a range of fetal and maternal complications that could impact the outcome of any concurrent infection.^1^ The first trimester bears a major risk for miscarriage and fetal developmental abnormalities. The late second and third trimesters carry an increased likelihood for the development of maternal conditions, such as gestational diabetes and hypertensive disorders, which contribute to maternal morbidity and premature birth.

It is well known that infectious pneumonia is a common cause of morbidity and mortality in pregnant women due to several physiological factors such as lower lung volumes and increased oxygen consumption.^2,3^ Indeed, a quarter of pregnancies complicated by pneumonia are estimated to require critical care hospitalization and ventilation support.^4^ Pregnancy-induced changes to immunity, such as reduction in cell-mediated cytotoxicity, diminished lymphokine response and reduction in lymphocyte proliferative response, further add to the prognostic picture of pneumonia in pregnant women.^3^ Hence, the advent of the SARS-CoV-2 pandemic could raise new questions about perinatal and obstetric management. Here, we review 164 pregnancies reported in the literature across 20 studies.

## Methods

We searched PubMed using the terms “pregnancy,” “pregnant,” “prenatal,” “Covid-19,” “SARS-CoV2”, “miscarriage,” “fetal anomalies” and “complications.” We limited our search to all published studies between December 20th, 2019 to April 18th, 2020 and added a filter for English-language articles. Our search retrieved 84 papers. We carried out the same search on MedRxiv, which yielded 27 results. We excluded reviews, meta-reviews, letters to the editor, and guidelines that are specific to a region. We then reviewed the results for relevance and included the articles that reported on SARS-CoV-2 in pregnant patients. This search strategy resulted in 19 case reports and articles from the primary literature that we included in this review. We added one study outside the aforementioned date range due to its novelty and relevance to the subject at hand. We extracted all information about patient characteristics, symptoms, laboratory results, and imaging studies. We also gathered the relevant data about the pregnancy course such as delivery method, gestational complications, and neonatal outcomes.

## Results

Results from individual studies are described and summarized in Table 1 and Table 2 below.

**Table 1:** Patient Characteristics and Symptoms in SARS-CoV-2 positive pregnancies

| Study | Number of patients | Maternal age (years) | Gestational age (weeks) | Pre-existing comorbidities | Maternal symptoms prepartum | Maternal symptoms postpartum | Imaging (CXR, CT) | Laboratory findings |
| --- | --- | --- | --- | --- | --- | --- | --- | --- |
| Alzamora, 2020 | 1 | 41 | 33 | Admitted due to resp insufficiency. PMH included diabetes mellitus (DM), and history of C-sections | Fever, malaise, fatigue, worsening dyspnea, tachycardia, tachypnea, metabolic acidosis | - | No CXR abnormalities<br>CT: multiple bilateral consolidations and bilateral pleural effusion | Elevated CRP, elevated ferritin, elevated D dimer, pancytopenia, lymphopenia<br>Low Hb, WBCs, platelets, and creatinine. |
| Baud, 2020 | 1 | 28 | 19 | Obesity<br>Admitted due to severe uterine contractions and a 5-cm dilated cervix at 19 weeks | Fever, fatigue, myalgia, dry cough, pain with swallowing, diarrhea | - | Physical examination did not show signs of pneumonia |  |
| Breslin, 2020 | 2 | 38, 33 | 37 (both) | Patient 1 admitted for poorly-controlled type 2 DM and intrahepatic cholestasis of pregnancy | Negative for fever, cough, dyspnea, or sore throat | Patient 1: Fever<br>Patient 2: Cough that progressed to respiratory distress, fever, tachycardia, | Patient 2 CXR: mild pulmonary vascular congestion with no consolidation or effusion | - |
|  |  |  |  | Patient 2 admitted for labor induction due to worsening chronic hypertension. PMH included DM and asthma |  | 88% O2 saturation, dyspnea, diaphoresis |  |  |
| Chen H, 2020 | 9 | Mean 30 (Range 26-40) | 36-39 | Not remarkable | Fever (7), cough (4), myalgia (3), sore throat (2), and malaise (2), GI symptoms (1), dyspnea (1) | Fever (6) | CT: patchy ground glass shadows (8) | Elevated CRP (6), lymphopenia (5), and increased ALT and AST (3) |
| Chen S, 2020 | 5 | Mean 28.8 (Range 25-31) | 38-41 | - | Cough (2), runny nose (1) | Fever (5) | CT: ground glass opacities bilaterally (3) or unilaterally (2) | Elevated CRP, decreased albumin, elevated alkaline phosphatase |
| Fan, 2020 | 2 | 34, 29 | 36, 37 | - | Nasal congestion (2), fever (2), abdominal rash (1), sore throat (1), chills (1) | - | CT patient 1: patchy consolidation in both lungs<br>CT patient 2: multiple patchy infiltrates on left lung | Lymphopenia (2) |
| Gidlof, 2020 | 1 | 34 | 36 | Referred due to hypertension and proteinuria. Dichorionic twin pregnancy | Headache, hoarseness, malaise, high BP, photophobia, brisk patellar reflexes | 87% O2 saturation | CT: typical signs of SARS-CoV-2 pneumonia | Elevated uric acid |
| Karami, 2020 | 1 | 27 | 30+3 | None | Fever, cough, myalgia, Respiratory Distress | Multi Organ Failure and maternal death | CXR: faint bilateral patchy opacities<br>CT: faint subpleural ground-glass opacities with pleural thickening | Leukopenia, lymphopenia, Elevated CPR, creatinine, and LDH |
| Lee, 2020 | 1 | 28 | 36+2 | - | Fever, cough, sore throat. Nausea and low BP following spinal anesthesia | - | CXR: left lower/middle lobe consolidation and increased vascular marking<br>CT: multifocal peribronchial ground glass appearance and consolidation in left lower lobe | Normal CRP, elevated erythrocyte sedimentation rate, low Hb |
| Li N,<br>2020 | 16** | Mean:<br>30.9<br>(Range<br>26-37) | 33-40 | Hypertension,<br>hepatitis B<br>infection, ,<br>polycystic ovary<br>syndrome | Fever (4) | Fever (8) | Typical images of<br>pneumonia<br>bilaterally (10) and<br>unilaterally (7) | Leukopenia, low<br>neutrophils, low CRP,<br>and low ALT |
| Li Y,<br>2020 | 1 | 30 | 35 | - | Dry cough,<br>chills, or<br>dyspnea | - | CXR: scattered<br>multiple patchy<br>infiltrates in both<br>lungs | Slightly abnormal |
| Liu D,<br>2020 | 15 | 23-40<br>(mean<br>32) | 12-38 | Mitral and<br>tricuspid valve<br>replacement | Fever (13),<br>cough (9),<br>sore throat<br>(1), dyspnea<br>(1), myalgia<br>(3), fatigue<br>(4), diarrhea<br>(1) | Fever (1) | CT: ground-glass<br>opacity (GGO).<br>consolidations<br>reported with disease<br>progression | Lymphocytopenia (12),<br>elevated CRP (10) |
| Liu H,<br>2020 | 41*** | Mean:<br>30 | 22-40+5 | Hepatitis B (1) | Fever (16),<br>cough (15),<br>dyspnea (5),<br>fatigue(5),<br>loss of<br>appetite (3) | Fever (14) | CT: Mixed GGO<br>with consolidation | Elevated CRP (27),<br>lymphopenia (25),<br>leukocytosis (17),<br>elevated neutrophils (34) |
| Liu W,<br>2020 | 3 | 34, 34,<br>30 | 40, 38+4,<br>39+5 | Patient 1:<br>hypothyroidism | Fever (2),<br>cough (2) | - | CT: abnormal in all 3<br>patients | Leukocytosis (2),<br>elevated neutrophils (2),<br>lymphocytosis (1), low |

|  |  |  |  | and epiglottic cysts<br>Patient 2: none<br>Patient 3: gestational diabetes |  |  |  | eosinophils (1), low platelets (1) |
| --- | --- | --- | --- | --- | --- | --- | --- | --- |
| Study | Number of patients | Maternal age (years) | Gestational age (weeks) | Pre-existing comorbidities | Maternal symptoms prepartum | Maternal symptoms postpartum | Imaging (CXR, CT) | Laboratory findings |
| Liu Y, 2020 | 13 | Mean: 29.7 (Range 22-36) | 2 women less than 28 GA (25, 27).<br>11 in third trimester (32 - 36) | None | Fever (10), cough (2), fatigue (4), dyspnea (3), sore throat (1) | 1 patient had MODS including ARDS | - | - |
| Nie, 2020 | 33 | Mean: 30.5 (Range 24-36) | 3 women in 2nd trimester (17, 20, 26), 30 women in 3rd trimester | 15 women had pre-existing chronic disorders | Fever (21), dry cough(13), fatigue(7), dyspnea (7)<br>Less common: diarrhea, muscle ache, sore throat, chest pain, stomachache<br>4 asymptomatic | Fever (5) | CT: bilateral or unilateral pneumonia, GGO | - |
| Vlachodimitropoulou, 2020 | 2 | 40, 23 | 35+3, 35+2 | Patient 1: neutropenia, mild respiratory infections<br>Patient 2: not reported | Pt 1: fever, cough, tachycardia<br>Pt 2: fever, cough | - | Pt 1: normal CXR<br>Pt 2: not reported | Pt 1: progressive thrombocytopenia, declining fibrinogen, rising APTT, elevated CRP, low neutrophil count, lymphocytopenia, elevated D dimer<br>Pt 2: thrombocytopenia, prolonged APTT, transaminitis, progressive coagulopathy, elevated CRP, lymphocytopenia, elevated D dimer |
| Wen, 2020 | 1 | 31 | 30 | - | Mild diarrhea | Recovered and discharged prior to delivery | CT: patchy consolidation and GGO | Not reported |
| Yu, 2020 | 7 | Mean: 32 (Range 29 - 34) | 39+1 (37 - 41+2) | 2 patients had chronic diseases (hypothyroidism and polycystic ovary syndrome)<br>3 | Fever (6), cough (1), dyspnea (1), diarrhea (1) | - | CT: bilateral pneumonia (6), unilateral pneumonia (1) | Elevated CRP (7), elevated neutrophils (5), lymphopenia (5), low platelets (2), elevated D- dimer (7), abnormal liver enzymes (2) |
| Zhu, 2020 | 9 | Mean: 30<br>(Range 25-35) | 31-39 | - | Fever (8), cough (4), diarrhea (1), sore throat (1), cholecystitis (1) | - | CT: typical of viral pneumonia. Lesions merged into strips as disease progressed | - |
| **Total n** | **164** |  |  |  |  |  |  |  |

\*Mixed diagnosis: 25 clinically-diagnosed and 16 laboratory-diagnosed
\*\*Compared with non-SARS-CoV-2 pneumonia pregnant women
\*\*\*Compared with 14 non-pregnant patients
- in Table 1 is used to refer to information that was not reported in the publication

**Table 2:**
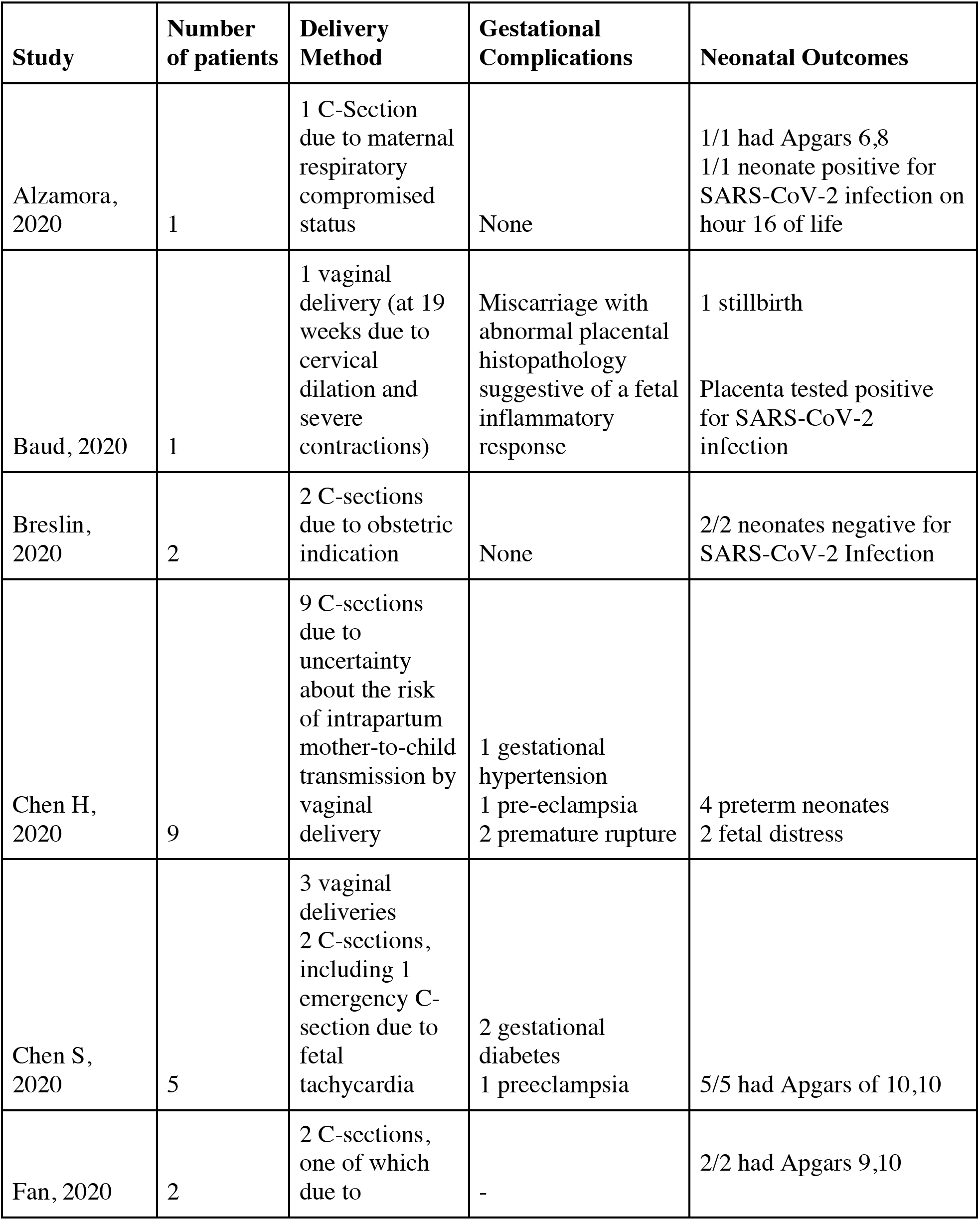
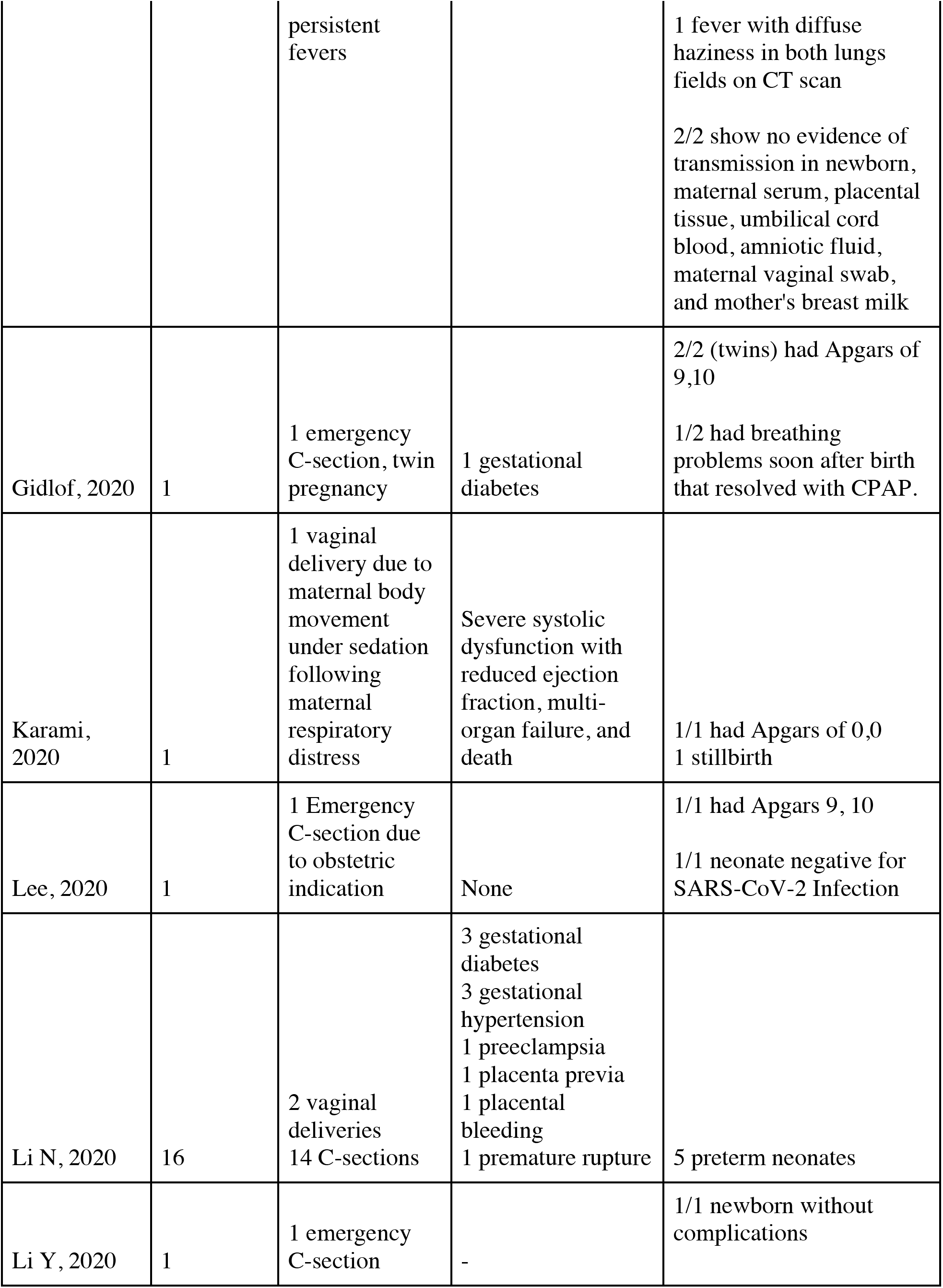
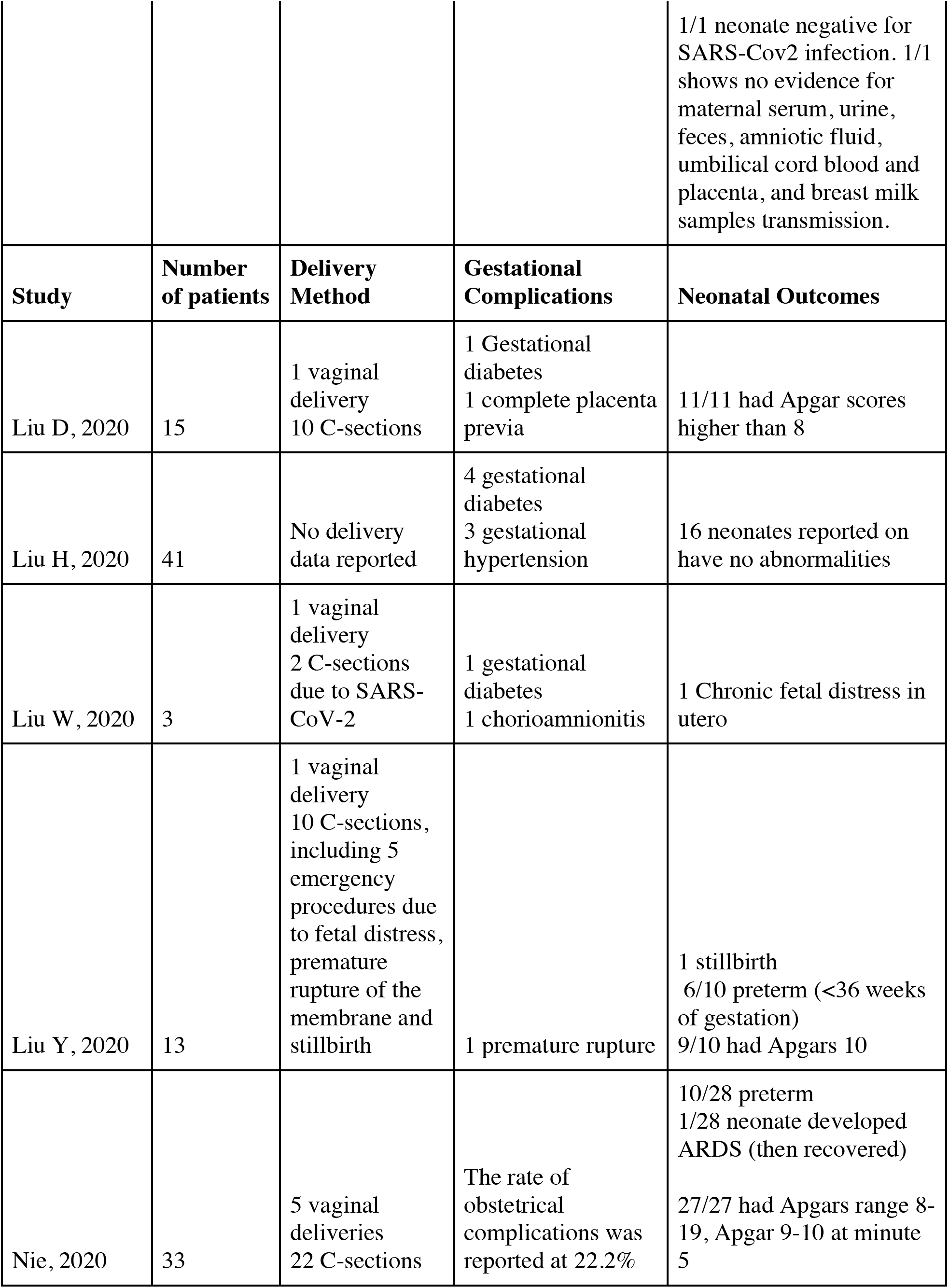
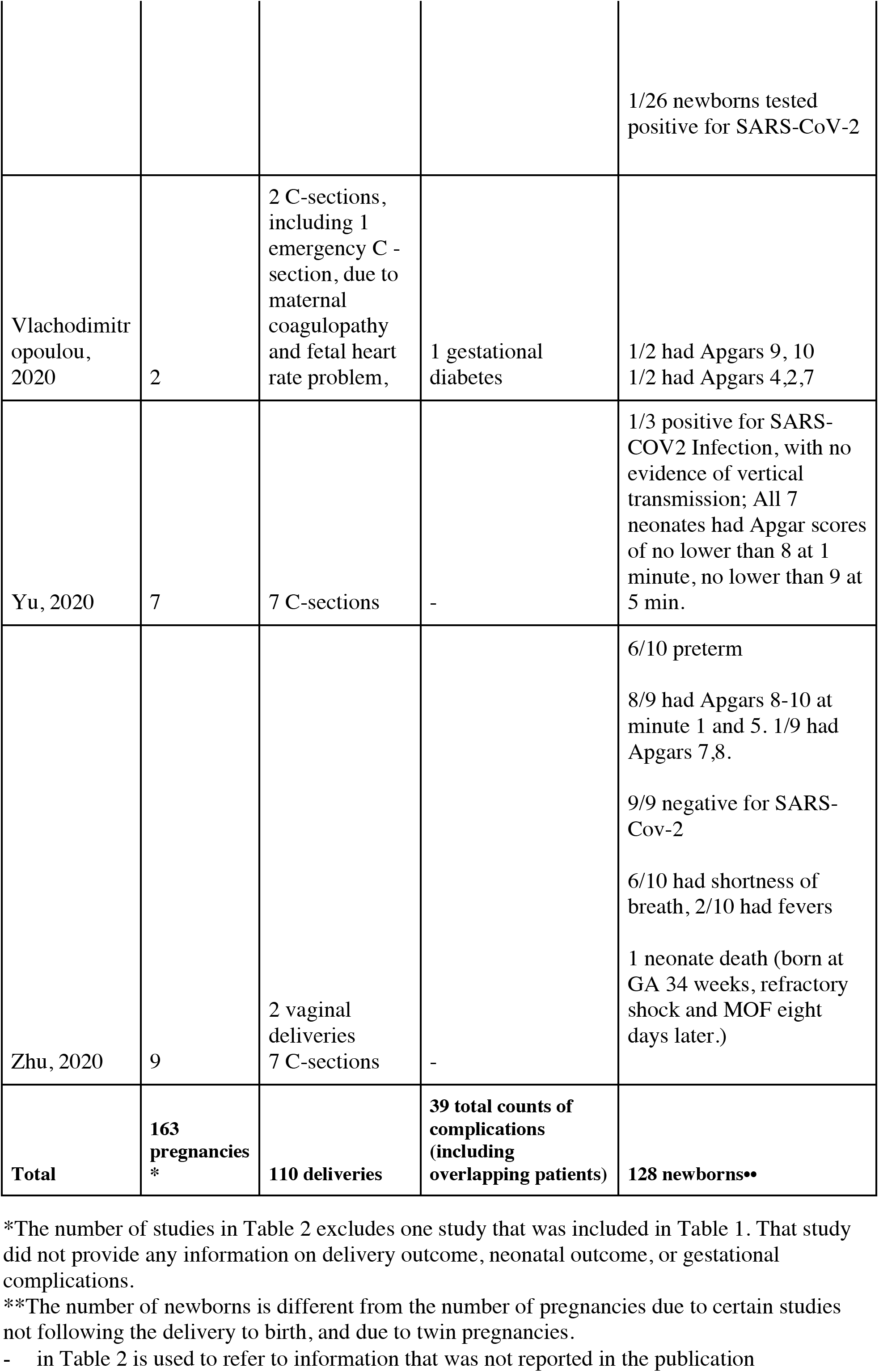
Summary of Gestational Complications and Delivery Outcomes in SARS-CoV-2 positive pregnancies

### Maternal Symptoms

With regards to prepartum symptoms, the most common symptoms were fever (57.9%) followed by cough (35.4%), fatigue (15.2%) and dyspnea (12.2%). Less common symptoms included diarrhea and gastrointestinal symptoms, sore throat, nasal congestion and runny nose, muscle aches, rash, headache, hoarseness, myalgia, high blood pressure, tachycardia, and cholecystitis. Only 4.3% of all patients were asymptomatic prior to delivery. Two patients who were asymptomatic prior to delivery became symptomatic post-partum, both requiring ICU admission.

The majority of the studies included patients in the third trimester only with the exception of four studies which also included patients in the second trimester. Most of these studies reported the number of patients experiencing each symptom without specifying the GA, so it is difficult to determine whether patients in earlier trimesters differed in their clinical presentations from those in later trimesters. D. Liu and colleagues (2020) reported 15 patients who ranged in GA from 12 to 38 weeks, but it was not clear whether the patients in the earlier trimester experienced different symptoms or complications. ^5^ Nie (2020) included 33 patients in total which ranged in GA from 24 to 36 weeks.^6^ No ICU admissions or mortalities were reported, and the study concluded that pregnant women were not at an increased risk for severe illness or mortality. Based on the findings reported by Y. Liu et al. (2020) and Nie (2020), patients’ clinical presentations do not appear to differ based on the gestational age of pregnancy.

Among the reported postpartum symptoms, the most common one was fever (24.7%). In one patient who was admitted with worsening hypertension and a history of diabetes mellitus and asthma, postpartum symptoms included low oxygen saturation, tachycardia, dyspnea, diaphoresis, and cough that progressed to respiratory failure.^7^ Gidlof et al. (2020) also reported low oxygen saturation postpartum. ^8^ Among 13 patients, Y. Liu and colleagues (2020) reported that one patient developed Multi Organ Dysfunction Syndrome (MODS) including Acute Respiratory Distress Syndrome (ARDS). ^9^ Karami et al. (2020) also reported a case of pre-partum respiratory distress, which progressed to multi-organ failure post-partum and unfortunately in the eventual death of the patient and the fetus.^10^ Thus, out of 164 pregnant women with SARS-CoV-2, 4 (2.44%) developed respiratory distress.

### Laboratory Findings

The most common laboratory abnormalities seen in pregnant women infected with SARS-CoV-2 were elevated C-reactive protein (CRP), elevated neutrophils, and leukocytosis. In the studies that reported laboratory findings, 114 pregnant women were included and among them 50.9% had elevated CRP, 44.7% had lymphopenia, 34% had elevated neutrophils, and 16.7% had leukocytosis. Among 24 pregnant women with SARS-CoV-2, Liu H reported that 58.6% of them had low or normal leukocyte levels.

Other laboratory abnormalities included low platelets, elevated D-dimer, abnormal liver enzymes, decreased albumin, low hemoglobin, elevated alkaline phosphate, decreased eosinophils, elevated uric acid, elevated ferritin, pancytopenia, prolonged APTT, low fibrinogen, and progressive coagulopathy.

### Chest X-ray (CXR)

Most studies reported imaging abnormalities in either chest X-ray (CXR), computed tomography (CT), or both. In terms of CXR, Y. Li et al. (2020) (n=1) reported scattered multiple patchy infiltrates in both lungs, Lee et al. (2020) (n=1) reported left lower/middle lobe consolidation and increased vascular marking, while Breslin et al. (2020) (n=1) reported mild pulmonary vascular congestion with no consolidation or effusion ^7,11,12^ All three women were of similar GA (mean 36 weeks).

### Computed Tomography (CT) Scan

Typical signs of viral pneumonia were frequently reported, with most patients having bilateral lung involvement. CT findings did not show signs of worsening pneumonia postpartum. However, with disease progression, Zhu et al. (2020) reported that lesions merged into strips and D. Liu et al. (2020) observed a paving pattern and consolidation. ^5,13^ Ground glass opacities (GGO) were reported in the majority of studies. Mixed GGO with consolidation were more common among pregnant SARS-CoV-2 patients in comparison to non-pregnant SARS-CoV-2 patients.^5^ Lee and colleagues (2020) also reported GGO with consolidation.^11^ In addition to multiple bilateral consolidations, bilateral pleural effusion were observed. ^14^ Given the prevalence of respiratory symptoms in pregnant women infected with SARS-CoV-2, lung ultrasound can a valuable tool in confirming diagnosis of SARS-CoV-2 infection as well as monitoring disease progression.^15^ Lung ultrasound eliminates the radiation risk and avoids the incidence of false-negative results associated with other diagnostic tests. As outlined by Moro and colleagues (2020), specific guidelines can be followed when performing lung ultrasound for pregnant women infected with SARS-CoV-2. ^15^

### Maternal and Obstetric Complications

There have been several reports of uncomplicated term delivery in SARS-CoV-2 positive women. ^11,16^ Of 110 deliveries reported, 93 were performed as C-sections, including 10 that were performed as emergency surgeries. The rate of C-sections is thus 84.5% of all deliveries, significantly higher than the expected rate of approximately 15% for pregnancies in the general population.^17,18^ This rate of C-section is also higher than that in pregnancies complicated by viral Influenza infection, where the likelihood of C-section did not increase relative to the general population.^19^

Several case reports highlight the frequency of C-section deliveries compared to vaginal deliveries, whether in healthy-appearing patients or patients presenting with fever.^13,20^ The indications for C-section varied and were not consistently reported across studies. In certain hospitals, the choice of C-section was determined by obstetric factors.^21^ In others, C-section was performed proactively due to maternal SARS-CoV-2 infection. Presumably, C-sections minimize cross-infection and reduce maternal exertion during labor, as per certain recommendations.^22^ Due to the uneven distribution of C-sections as opposed to vaginal delivery, it is not possible to comment on differences in neonatal and maternal outcomes between vaginal delivery and C-sections in SARS-CoV-2 pregnancies. Importantly, there are no contraindications for vaginal delivery as evidenced by the uncomplicated vaginal delivery in at least five patients.^23 24^ It is not clear from this review if the high Cesarean section rate is warranted.

Out of 163 pregnancies reviewed, we looked at the following gestational complications: gestational diabetes, preeclampsia or gestational hypertension, and placental complications such as premature rupture or placenta previa. The rate of complications was at 23.93%, composed of gestational diabetes (7.99%), preeclampsia or gestational hypertension (5.52%), and placental complications (6.13%). At least 6 additional patients (3.68%) had complications, but it is not possible to ascertain the specific condition due to the method of reporting in the primary literature.

### Fetal and Neonatal Outcomes

The 110 pregnancies followed in the literature resulted in 128 newborns. Out of 128 neonates, 26 (20.31%) were delivered preterm, before 36 weeks of gestation Again, it is not consistently clear whether early delivery was induced in light of obstetric indications or maternal SARS-CoV-2 infection. The vast majority of neonates for whom an Apgar score was reported (n=68) scored in the normal range, at least 8 at minute 1 and minute 5, with the exception of three neonates. Three newborns (4.41%) had a concerning Apgar score of <7 at minute 1.

Eight fetuses (6.35%) exhibited fetal distress, which was the indication for preterm delivery in some cases. Postnatally, the rate of respiratory distress or abnormal lung findings among the neonates was at 7.94%, divided as follows: 6 neonates had shortness of birth, 2 had pneumonia-like presentation on imaging, and two had respiratory distress including one case of ARDS. All cases resolved with antibiotics without complication. Additionally, three newborns presented with fever (2.34%).

Of the 128 fetuses, four had very poor outcomes: three deliveries resulted in stillbirth, including one fetus who was born to a mother who passed away soon after delivery. ^9,10,25^ One neonate developed refractory shock and multiple organ failure and passed on day 8 of life.^13^ Although no fetus is thought to have died as a result of fetal infection, Baud et al. (2020) suggest that placental inflammation in the setting of the virus contributes to fetal distress, even when the fetal is not infected.^25^ Notably, several case studies have ruled out vertical transmission of SARS-CoV-2. This is largely supported by our review. Out of the neonates tested for SARS-CoV-2 infection (n=44), 3 tested positive for the infection through nasopharyngeal swab (6.81%). Two of the cases in question raised concerns about potential vertical transmission, since the mother and the obstetric team practiced protection and isolation measures to prevent infection during the delivery process and immediately afterwards.^6,14^ However, because of the asymptomatic presentation of many infected individuals, it is possible that the neonates had acquired SARS-Cov-2 in the hospital after birth.

Certain studies went beyond testing the newborn for evidence of vertical transmission. Gidlof et al. (2020) tested breast milk and maternal vaginal secretion of a SARS-CoV-2 positive mother.^8^ Y. Li et al. (2020) tested the breast milk, amniotic fluid, umbilical cord blood and placenta, maternal serum, urine, and feces of a SARS-CoV-2 positive mother. ^9^ Fan (2020) tested breast milk, maternal vaginal secretion, placental tissue, umbilical cord blood, amniotic fluid and maternal serum in two pregnancies.^20^

### Breastfeeding

Collected breast milk samples from the four aforementioned pregnancies were negative for SARS-CoV-2. These results, in addition to several guidelines such as those published by the ACOG and the RCOG, converge on the idea that SARS-CoV-2 is not carried through breast milk.^26,27^ However, the underlying risk of neonatal exposure to maternal respiratory droplets during the breastfeeding process persists. Overall, the many benefits of breastfeeding do make breastfeeding safe, as long as SARS-CoV-2 positive mothers take reasonable precautions while in close contact with the newborn.

## Discussion

According to Liu et al., pregnancy and delivery did not aggravate the severity of SARS-CoV-2 in the patients studied. ^5^ However, as one expects, the variability in the severity of infection and pneumonia presentation in different patients makes such a statement difficult to prove.

Pregnancy, in particular the third trimester, is associated with changes in lung physiology, such as decreases in ERV and FRC volumes and increases in respiratory resistance.^28^ A prime example is the fact that pregnant women infected with the H1N1 virus during the 2009 epidemic were at a significantly higher risk of complications and hospitalization than non-pregnant women. ^29,30^ Hence, an increased rate of respiratory complications, in addition to maternal and neonatal complications, could be expected in pregnant patients with SARS-CoV-2 infection. The absence of widespread respiratory complications is surprising and suggests that a larger number of patients will help better understand the natural history of SARS-CoV-2 infections during pregnancy.

It is certainly true that, at least from the available reports, that the mortality risk in SARS-CoV-2 pregnant women is extremely low. There exists only one case of maternal mortality and three other cases of patients developing respiratory distress. ^7–9^ At the time of writing, there exists a single report of a (late) miscarriage due to SARS-CoV-2 infection, and no reports of termination due to early congenital defects. In the 2002-2004 SARS epidemic, miscarriage rates were reported to be as high as 50% for infected women in the first trimester.^31^ Similarly, infection with the seasonal influenza virus is associated with higher miscarriage and maternal mortality rates than the noninfected population.^1,29^ It should be noted however that miscarriage could be due to the maternal response to the infection, rather than a direct placental effect induced by the infection.^32^ In the case of SARS-CoV-2, it is highly likely that the higher rate of miscarriage could be attributed to fever. In contrast to these viral infections, the outcomes for SARS-CoV-2 infected pregnant women in the first trimester are reassuring.

With regards to fetal and neonatal outcomes, pneumonias during pregnancies are associated with an increased risk of preterm birth compared to the general population.^33^ Our findings are consistent with this observation. Fortunately, premature neonates fared well and had comparable Apgar scores to the full-term neonates included in the analysis. Unfortunately, the rate of stillbirth among this group was 2.34%, significantly higher than the rate among the general population. Intrauterine growth restriction (IUGR) was an expected finding in this group due to its prevalence in prior SARS infections during pregnancy.^34^ However, there was little reporting on IUGR among fetuses in SARS-CoV-2 positive pregnancies.

Although our review includes a high number of patients, it is limited by the heterogeneity of the studies’ primary outcomes and reporting methods. Secondly, the availability of few reports of severe outcomes makes it difficult to explain the pathophysiology of stillbirths and maternal morbidity in the setting of SARS-CoV-2. As more case reports become available, sub-group analyses will enable stronger understanding of the impact of the virus on maternal and fetal health.

## Data Availability

As this is a systematic review, all the data is available form the articles included in the review and listed in the references section.

